# Very early neurological deterioration after thrombolysis in patients with acute ischemic stroke

**DOI:** 10.1101/2023.03.05.23286824

**Authors:** Ying-Chi Shen, Shin-Joe Yeh, Chih-Hao Chen, Sung-Chun Tang, Li-Kai Tsai, Jiann-Shing Jeng

## Abstract

**Background:** Early neurological deterioration within 24 h after thrombolysis in patients with acute ischemic stroke (AIS) is associated with poor outcomes. Evidence is lacking regarding neurological deterioration within 1 h after thrombolysis.

**Methods:** Patients who received intravenous thrombolysis with tissue plasminogen activator (tPA) for AIS between January 2018 and December 2021 were consecutively enrolled. Very early neurological deterioration (VEND) was defined as a ≥ 4-point increase in the National Institutes of Health Stroke Scale (NIHSS) score within 1 h after starting thrombolysis compared with the pre-treatment score. A modified Rankin Scale score of 3–6 at 3 months was defined as a poor functional outcome.

**Results:** Of the 353 AIS patients (age 69.7 ± 13.3 years, 57% men) receiving thrombolysis with tPA, 29 (8.4%) had VEND. VEND was associated with symptomatic intracranial atherosclerotic disease (ICAD) (41% vs. 17%, *P* = 0.005) and was an independent predictor of poor functional outcomes at 3 months (adjusted odds ratio 3.04, *P* = 0.043). The VEND group had higher NIHSS scores at 1 h (19.2 ± 7.3 vs. 9.0 ± 7.1, *P* < 0.001) and 24 h (14.1 ± 9.8 vs. 7.3 ± 7.5, *P* = 0.001) after initiating tPA than the non-VEND group. In patients with an initial NIHSS score < 6, VEND was significantly associated with ICAD, receiving endovascular thrombectomy (EVT), and poor functional outcomes. In patients with VEND, EVT with successful recanalization led to lower NIHSS scores at 24 h than in those without successful recanalization (12 ± 9 vs. 26 ± 7, *P* = 0.047), and 24-h NIHSS scores predicted poor functional outcomes.

**Conclusions:** In patients receiving thrombolysis, VEND was independently associated with poor functional outcomes. Identifying VEND is crucial for underlying ICAD and salvageability by EVT. Successful recanalization by EVT effectively reduced 24-h stroke severity in patients with VEND.

## Introduction

Intravenous thrombolysis with tissue plasminogen activator (tPA) has been shown to improve functional outcomes in patients with acute ischemic stroke (AIS) within 4.5 h.^1^ Certain patients exhibit neurological deterioration within the subsequent 24 h after receiving tPA, which is referred to as early neurological deterioration (END).^2^ END is usually defined as an increase of ≥ 4 points in the National Institutes of Health Stroke Scale (NIHSS) score within 24 h compared to the baseline score.^3^ The incidence of END after thrombolysis among patient with AIS is 10–30%.^3-5^ END is also consistently associated with poor long-term outcomes with increased risk of death and dependency.^6-8^

Although several mechanisms of END after thrombolysis have been revealed, such as early recurrent ischemic stroke, symptomatic intracranial hemorrhage (sICH), and malignant cerebral edema, two-thirds of patients with END after thrombolysis lack an identifiable cause.^6,9^ Among these etiologies of END, ischemic progression is approximately twice as common as sICH or early brain edema.^5,6^ In addition, ischemic mechanism is more predominant in the early hours than other ones after thrombolysis.^10,11^ Ischemic-type END was reported to occur at a median of 4 h after tPA, while hemorrhagic-type was at 8 h.^11^ Moreover, detection of neurological deterioration within 1 h after initiating tPA is crucial because it is within 6 h of stroke onset, fulfilling the criteria for endovascular thrombectomy (EVT) if large-vessel occlusion is further confirmed.^1^ Since neurological assessment every 15 minutes within the first 2 hours after initiation of tPA is a routine practice,^12^ neurological deterioration within the first hour can be identified. Thus, we defined neurological deterioration within 1 h after initiating tPA as very early neurological deterioration (VEND).

No prior study has explored the etiology and impact of VEND after tPA treatment, and the benefit of EVT for VEND. Hence, we aimed to investigate the prevalence, predictive factors, influence of VEND after thrombolysis, and the effect of rescue EVT for VEND in patients with AIS.

## Methods

### Study design and patient enrollment

We performed a retrospective analysis of the prospectively collected data from the National Taiwan University Hospital Stroke Registry initiated in 1995, which was approved by the Research Ethics Committee Office (No. 9561703056). All patients with AIS who received thrombolysis at this hospital or being transferred to this hospital immediately after initiating thrombolysis between January 2018 and December 2021 were consecutively included in this study. Thrombolytic therapy was administered according to treatment guideline.^1^ The standard-dose of tPA was 0.9 mg/kg,^1^ while low-dose (0.6 mg/kg) could be applied in patients older than 70 years old according to the recommendation of the Taiwan Food and Drug Administration.^13,14^ In the total dose of tPA, 10% was administered as a bolus within 4.5 h of symptom onset, followed by a continuous intravenous infusion of the remaining dose for 1 h.^15^ Neurological evaluation was performed every 15 min in the first 2 h after starting thrombolysis according to guideline.^12^ Detailed NIHSS scores were assessed immediately before thrombolysis, at 1 and 24 h after initiating thrombolysis, and at any time when deterioration was noticed.^12^ For patients who were receiving EVT at 1 h after initiating thrombolysis, abbreviated neurological evaluation could be performed during EVT procedure to monitor any neurological deterioration, and a NIHSS score was routinely assessed after completion of EVT. Most patients underwent EVT with consciousness sedation, while general anesthesia was applied for uncooperative or intubated patients.^16^ Patients who lacked initial NIHSS scores, initial laboratory data, or brain image at 24 h after thrombolysis, or who were treated with tenecteplase, were excluded from this study.

### Data collection

Demographics (age, sex), vascular risk factors (hypertension, diabetes mellitus, dyslipidemia, heart disease, chronic kidney disease, prior stroke, cancer, smoking status, and alcohol consumption), and prior use of antiplatelet or anticoagulant therapy were retrieved from medical records. We retrieved the NIHSS scores immediately preceding thrombolysis, at 1 h and 24 h after initiating thrombolysis, and when neurological deterioration was identified. VEND was defined as an increase of ≥ 4 points in the NIHSS score within 1 h of the initiation of thrombolysis compared with the score immediately before thrombolysis. The relevant laboratory data on admission including platelet count, international normalized ratio (INR), creatinine, random blood glucose at admission, hemoglobin A1C (HbA1c), low-density lipoprotein cholesterol (LDL), triglycerides (TG), and d-dimer levels were recorded. Since changes in blood pressure before tPA might be associated with neurological deterioration,^5^ the blood pressures at initial presentation and before thrombolysis were collected. Occluded arteries on pretreatment computed tomography angiography (CTA) images were recorded. The etiologies of ischemic stroke were classified based on the International Trial of Org 10172 in Acute Stroke Treatment (TOAST) classification.^17^ Large-artery atherosclerosis (LAA) etiology includes intracranial atherosclerotic disease (ICAD) and extracranial atherosclerotic disease (ECAD).^8,9^ LAA and symptomatic ICAD or ECAD were defined as ≥ 50% luminal stenosis or occlusion of the intracranial or extracranial artery contributing to this stroke, which was confirmed by vessel images after recanalization therapy.^18^ The parameters for thrombolysis included tPA dosages, door-to-needle time, and onset-to-needle time. The parameters obtained for EVT included the use of tirofiban and the modified Thrombolysis in Cerebral Infarction (mTICI) score at the end of EVT. The sICH complication was defined as clinical deterioration with corresponding intracranial hemorrhage within 7 days after thrombolysis.^19^ Functional outcomes were assessed 3 months after the stroke using the modified Rankin Scale (mRS), in which a poor functional outcome was defined as an mRS score of 3–6.^20,21^

### Statistical analysis

Continuous variables were presented as mean ± standard deviation and were compared using Mann–Whitney U test between the two groups. Categorical variables were presented as numbers (percentages) and were compared using Fisher’s exact test. Logistic regression analysis was used to investigate factors associated with poor functional outcomes and VEND after thrombolysis, respectively. Significant variables in the univariate models for predicting poor functional outcomes were further analyzed using multivariate logistic regression models. In patients with minor ischemic stroke (initial NIHSS score < 6) who were not considered candidates for EVT at initial presentation,^1^ the impact of VEND on functional outcomes was also evaluated. Lastly, we evaluated whether adding VEND improved the regression model composed of significant clinical variables for predicting poor functional outcomes at 3 months by comparing the areas under the receiver operating characteristic (ROC) curves. Statistical significance was set as *P* < 0.05. The Small STATA software (StataCorp LLC, College Station, TX, USA) was used for statistical analyses.

## Results

A total of 353 patients with AIS underwent intravenous thrombolysis during the study period. After excluding six patients with incomplete data and two patients treated with tenecteplase, data from the remaining 345 patients (mean age 69.7 ± 13.3 years, 57% men) were analyzed, in whom 29 (8.4%) had VEND (Supplemental Figure 1).

The clinical characteristics of the patients, according to the presence or absence of VEND, are summarized in Table 1. Compared to patients without VEND, those with VEND were more likely to have symptomatic ICAD (41% vs. 17%, *P* = 0.005), higher NIHSS scores at 1 h (19.2 ± 7.3 vs. 9.0 ± 7.1, *P* < 0.001) and 24 h (14.1 ± 9.8 vs. 7.3 ± 7.5, *P* = 0.001) after initiation of thrombolysis, and a higher probability of receiving EVT (59% vs. 25%, *P* < 0.001).

**Table 1.**
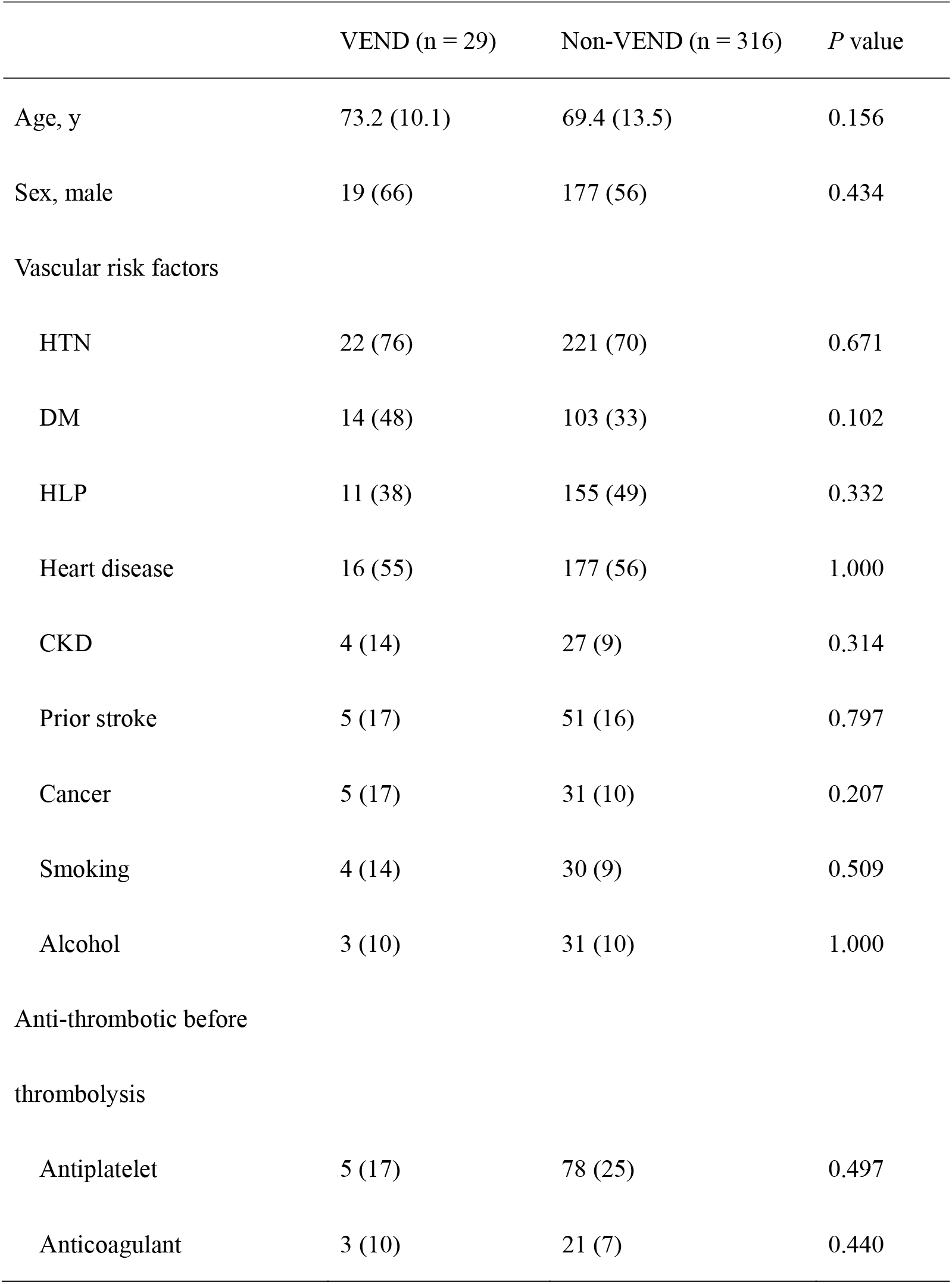
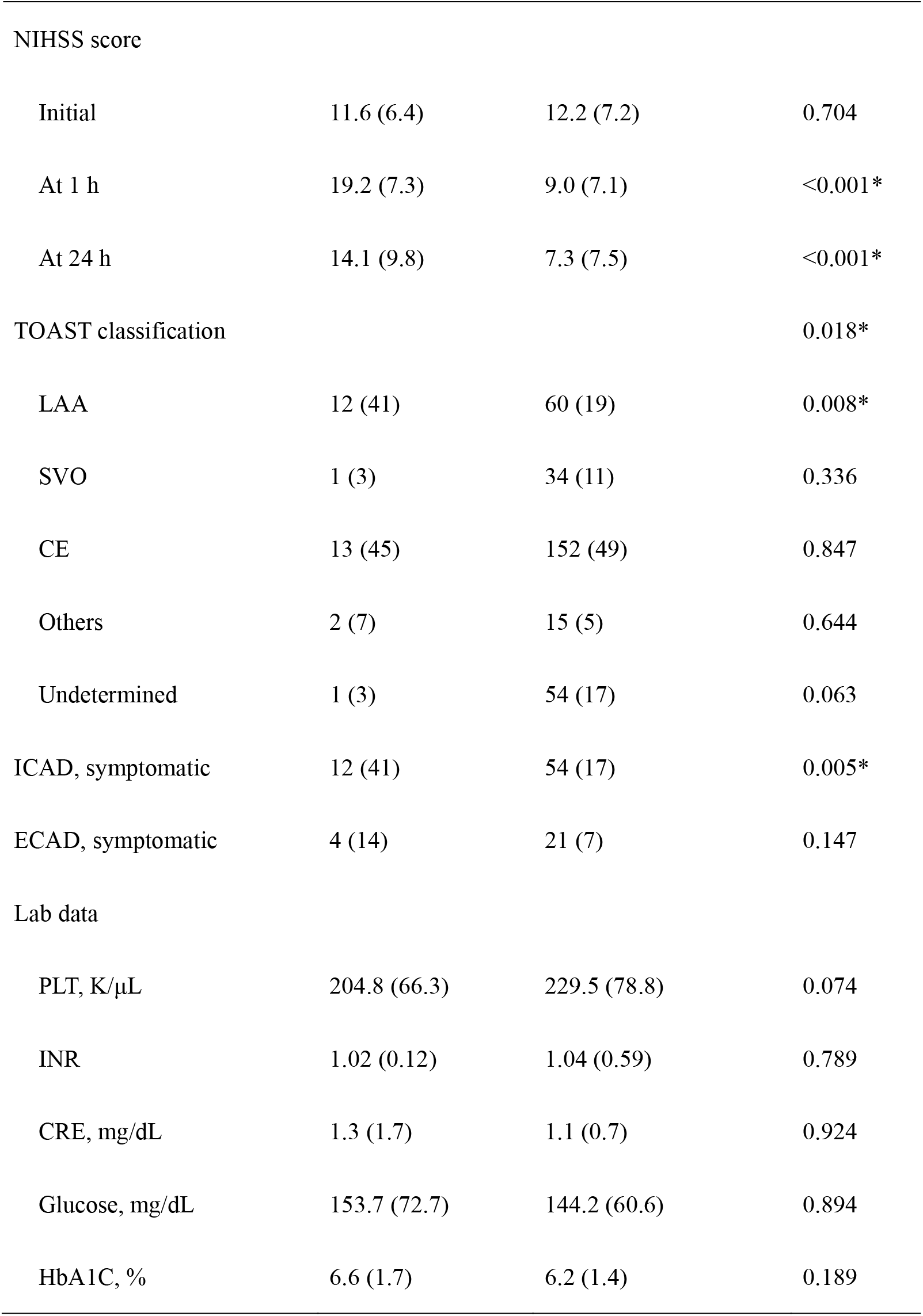
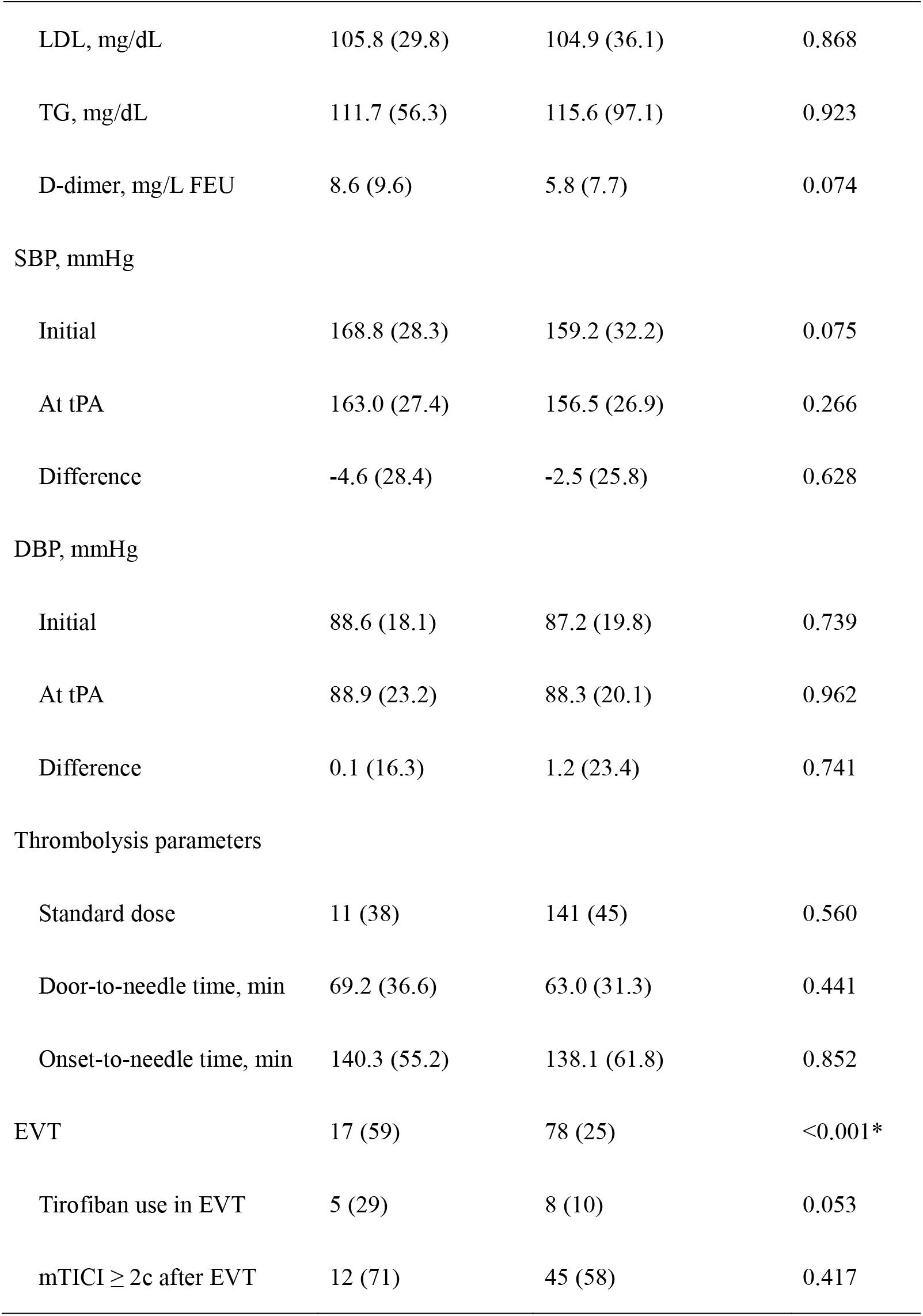
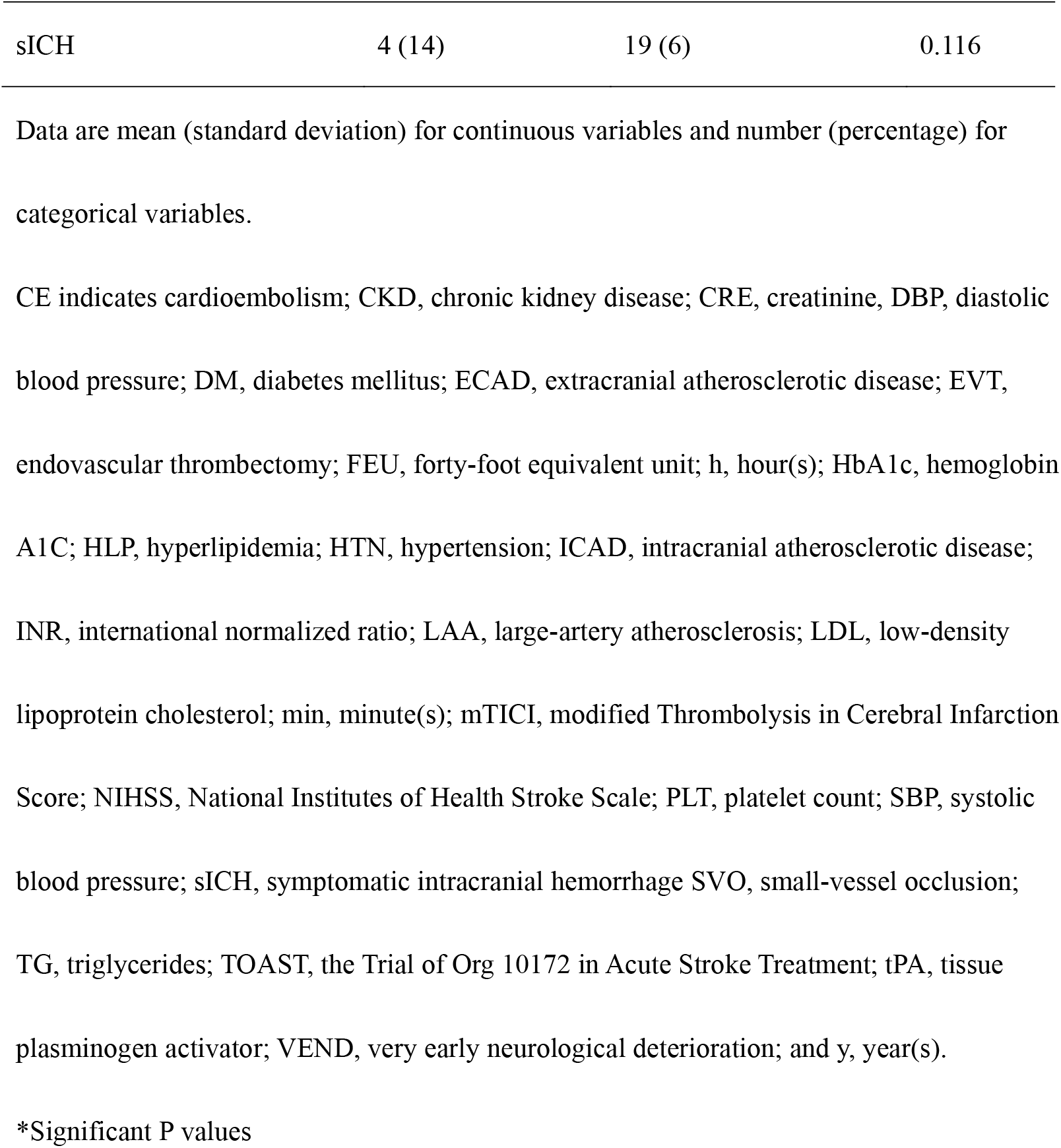
Characteristics of the patients with or without VEND after thrombolysis for acute ischemic stroke

|  | VEND (n = 29) | Non-VEND (n = 316) | *P* value |
| --- | --- | --- | --- |
| Age, y | 73.2 (10.1) | 69.4 (13.5) | 0.156 |
| Sex, male | 19 (66) | 177 (56) | 0.434 |
| Vascular risk factors |  |  |  |
| HTN | 22 (76) | 221 (70) | 0.671 |
| DM | 14 (48) | 103 (33) | 0.102 |
| HLP | 11 (38) | 155 (49) | 0.332 |
| Heart disease | 16 (55) | 177 (56) | 1.000 |
| CKD | 4 (14) | 27 (9) | 0.314 |
| Prior stroke | 5 (17) | 51 (16) | 0.797 |
| Cancer | 5 (17) | 31 (10) | 0.207 |
| Smoking | 4 (14) | 30 (9) | 0.509 |
| Alcohol | 3 (10) | 31 (10) | 1.000 |
| Anti-thrombotic before thrombolysis |  |  |  |
| Antiplatelet | 5 (17) | 78 (25) | 0.497 |
| Anticoagulant | 3 (10) | 21 (7) | 0.440 |
| NIHSS score |  |  |  |
| Initial | 11.6 (6.4) | 12.2 (7.2) | 0.704 |
| At 1 h | 19.2 (7.3) | 9.0 (7.1) | <0.001* |
| At 24 h | 14.1 (9.8) | 7.3 (7.5) | <0.001* |
| TOAST classification |  |  | 0.018* |
| LAA | 12 (41) | 60 (19) | 0.008* |
| SVO | 1 (3) | 34 (11) | 0.336 |
| CE | 13 (45) | 152 (49) | 0.847 |
| Others | 2 (7) | 15 (5) | 0.644 |
| Undetermined | 1 (3) | 54 (17) | 0.063 |
| ICAD, symptomatic | 12 (41) | 54 (17) | 0.005* |
| ECAD, symptomatic | 4 (14) | 21 (7) | 0.147 |
| Lab data |  |  |  |
| PLT, K/μL | 204.8 (66.3) | 229.5 (78.8) | 0.074 |
| INR | 1.02 (0.12) | 1.04 (0.59) | 0.789 |
| CRE, mg/dL | 1.3 (1.7) | 1.1 (0.7) | 0.924 |
| Glucose, mg/dL | 153.7 (72.7) | 144.2 (60.6) | 0.894 |
| HbA1C, % | 6.6 (1.7) | 6.2 (1.4) | 0.189 |
| LDL, mg/dL | 105.8 (29.8) | 104.9 (36.1) | 0.868 |
| TG, mg/dL | 111.7 (56.3) | 115.6 (97.1) | 0.923 |
| D-dimer, mg/L FEU | 8.6 (9.6) | 5.8 (7.7) | 0.074 |
| SBP, mmHg |  |  |  |
| Initial | 168.8 (28.3) | 159.2 (32.2) | 0.075 |
| At tPA | 163.0 (27.4) | 156.5 (26.9) | 0.266 |
| Difference | -4.6 (28.4) | -2.5 (25.8) | 0.628 |
| DBP, mmHg |  |  |  |
| Initial | 88.6 (18.1) | 87.2 (19.8) | 0.739 |
| At tPA | 88.9 (23.2) | 88.3 (20.1) | 0.962 |
| Difference | 0.1 (16.3) | 1.2 (23.4) | 0.741 |
| Thrombolysis parameters |  |  |  |
| Standard dose | 11 (38) | 141 (45) | 0.560 |
| Door-to-needle time, min | 69.2 (36.6) | 63.0 (31.3) | 0.441 |
| Onset-to-needle time, min | 140.3 (55.2) | 138.1 (61.8) | 0.852 |
| EVT | 17 (59) | 78 (25) | <0.001* |
| Tirofiban use in EVT | 5 (29) | 8 (10) | 0.053 |
| mTICI ≥ 2c after EVT | 12 (71) | 45 (58) | 0.417 |
| sICH | 4 (14) | 19 (6) | 0.116 |

Data are mean (standard deviation) for continuous variables and number (percentage) for categorical variables.
CE indicates cardioembolism; CKD, chronic kidney disease; CRE, creatinine, DBP, diastolic blood pressure; DM, diabetes mellitus; ECAD, extracranial atherosclerotic disease; EVT, endovascular thrombectomy; FEU, forty-foot equivalent unit; h, hour(s); HbA1c, hemoglobin A1C; HLP, hyperlipidemia; HTN, hypertension; ICAD, intracranial atherosclerotic disease; INR, international normalized ratio; LAA, large-artery atherosclerosis; LDL, low-density lipoprotein cholesterol; min, minute(s); mTICI, modified Thrombolysis in Cerebral Infarction Score; NIHSS, National Institutes of Health Stroke Scale; PLT, platelet count; SBP, systolic blood pressure; sICH, symptomatic intracranial hemorrhage SVO, small-vessel occlusion; TG, triglycerides; TOAST, the Trial of Org 10172 in Acute Stroke Treatment; tPA, tissue plasminogen activator; VEND, very early neurological deterioration; and y, year(s).
\*Significant P values

Considering that receiving EVT within 1 h after thrombolysis might impede the recognition of VEND, we found that the proportion of VEND in patients who received EVT within 1 h after thrombolysis did not differ from those after 1 h (13% vs. 8%, *P* = 0.266). Additionally, most of our patients (96%) underwent EVT with conscious sedation, which enabled physicians to detect neurological deterioration immediately during EVT procedure.

### Determinants of poor functional outcomes at 3 months

A total of 342 patients (99.1%) were followed for 3 months, in whom 150 (44%) had poor functional outcomes at 3 months. After adjusting for significant variables in logistic regression models, VEND remained a significant predictor of poor functional outcomes in patients with AIS receiving thrombolysis (adjusted odds ratio [aOR] 3.04, 95% confidence interval [CI] 1.04–8.90, *P* = 0.043) (Supplemental Table 1).

In patients with an initial NIHSS score of < 6 (minor stroke), VEND was the only significant predictor of poor functional outcomes at 3 months (odds ratio [OR] 8.00, 95% CI 1.64–39.04, *P* = 0.01) (Table 2). In patients with an NIHSS score of ≥ 6 (non-minor stroke), VEND was also an independent predictor of poor functional outcomes (OR 4.93, 95% CI 1.61–15.05, *P* = 0.005), along with age, initial systolic blood pressure (SBP), D-dimer, and mTICI ≥ 2c. After adjusting for age, VEND remained significantly associated with poor functional outcomes in both the minor and non-minor ischemic stroke groups (*P* = 0.010 and 0.012, respectively).

**Table 2.**
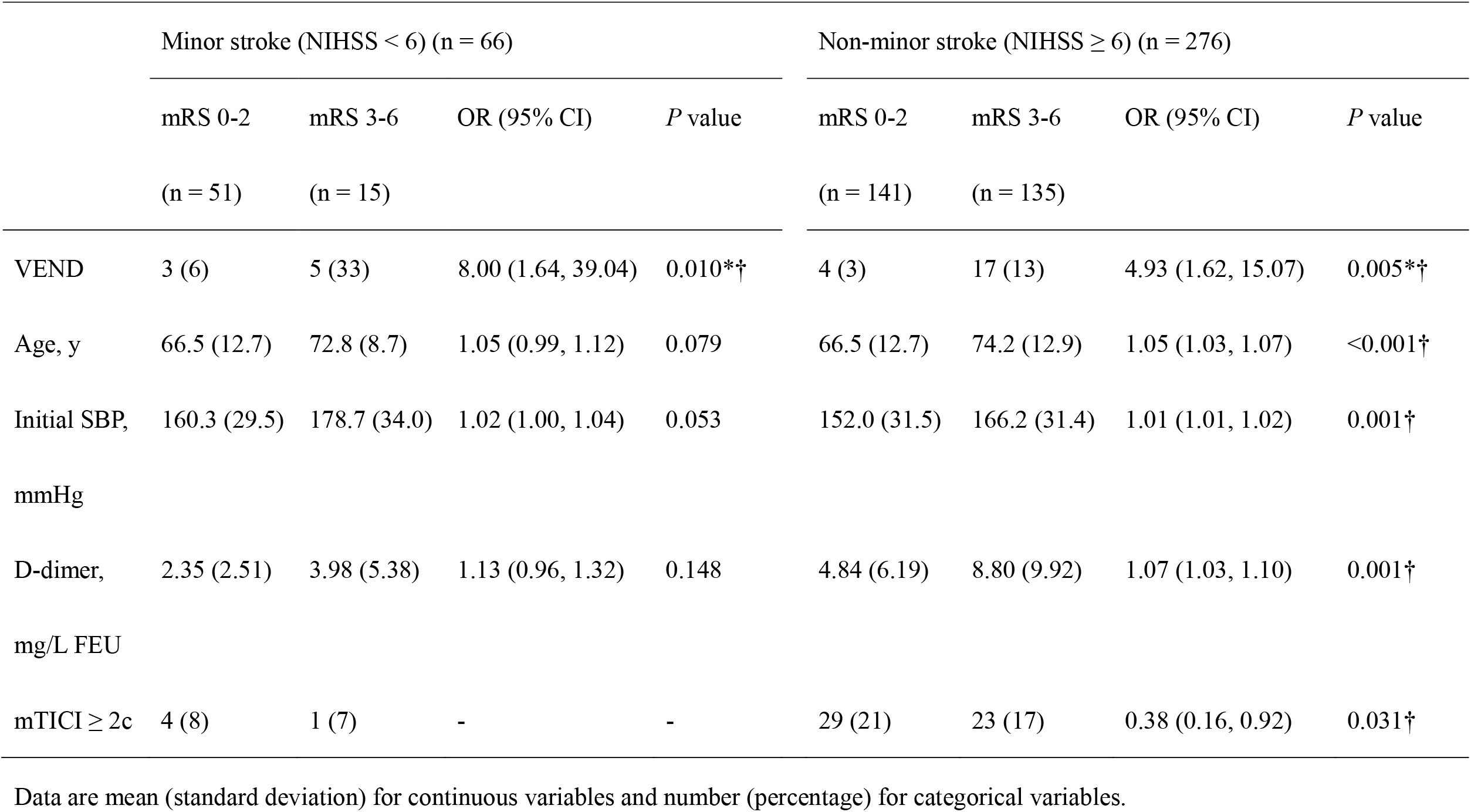
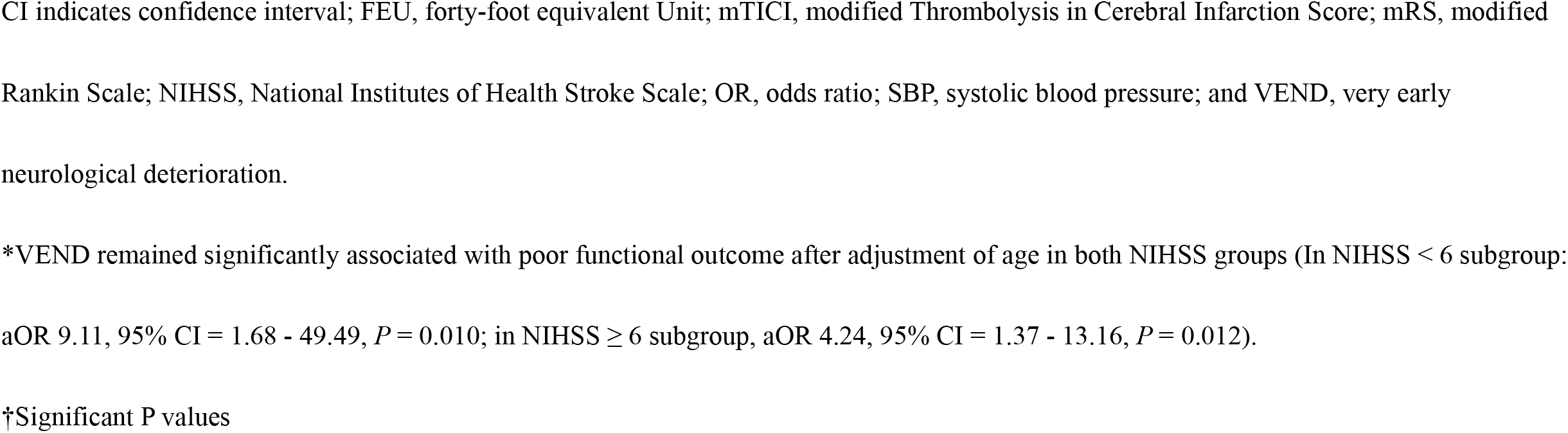
Factors associated with poor functional outcome in patients with minor and non-minor ischemic stroke

### ROC curve analysis for predicting long-term functional outcomes

To further clarify the predictive role of VEND in poor functional outcomes in patients with AIS receiving thrombolysis, a ROC curve analysis was applied to assess whether adding VEND in the reference model improved the predictive efficacy (Figure 1). Basic model A was composed of age, initial NIHSS score, and initial SBP, in which the area under the ROC curve (AUC) was 0.775 (*P* < 0.0001). Adding VEND to model A to produce model B (AUC = 0.802, *P* < 0.0001) significantly increased the AUC for prediction of poor functional outcomes (*P* = 0.047 for comparison between the two models).

**Figure 1.**
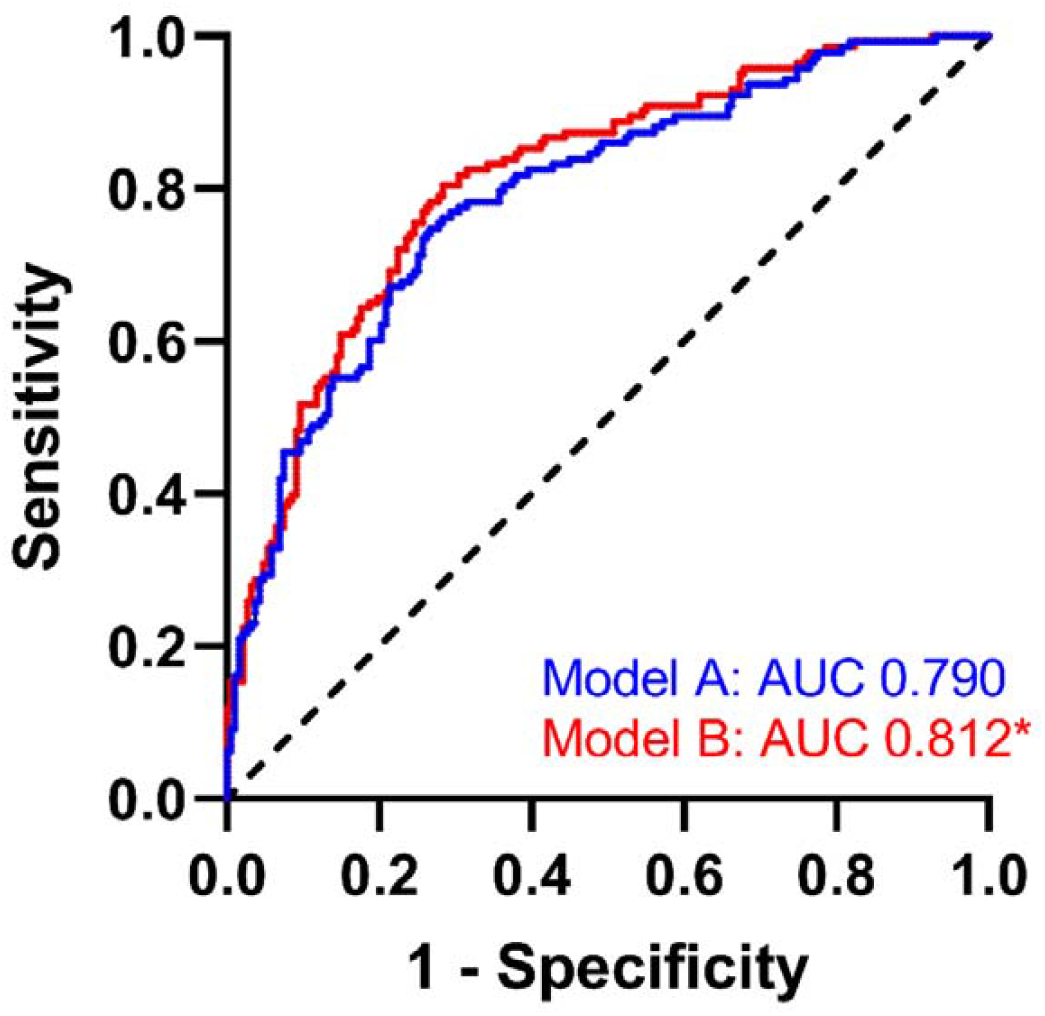
Receiver operating characteristic (ROC) curve analysis for prediction of poor functional outcomes. Model A was composed of age, initial NIHSS score, and initial SBP for predicting poor functional outcomes (AUC = 0.790, *P* < 0.0001). Adding VEND into model A to make model B (AUC = 0.812, *P* < 0.0001) significantly increased the AUC for prediction of poor functional outcomes (*P* = 0.048 for comparison of the two models). AUC, area under the ROC curve; NIHSS, National Institutes of Health Stroke Scale; SBP, systolic blood pressure; VEND, very early neurological deterioration.

### Risk factors for VEND

In patients receiving thrombolysis, the occurrence of VEND was associated with LAA etiology (OR 3.01, 95% CI 1.37–6.64, *P* = 0.006) (Table 3). LAA included symptomatic ICAD and ECAD, but only symptomatic ICAD (OR 3.42, 95% CI 1.55–7.58, *P* = 0.002) was associated with VEND. In terms of pretreatment occluded arteries, occlusion in the distal or proximal internal carotid artery (ICA) was significantly associated with VEND (*P* = 0.007 and 0.004, respectively).

**Table 3.**
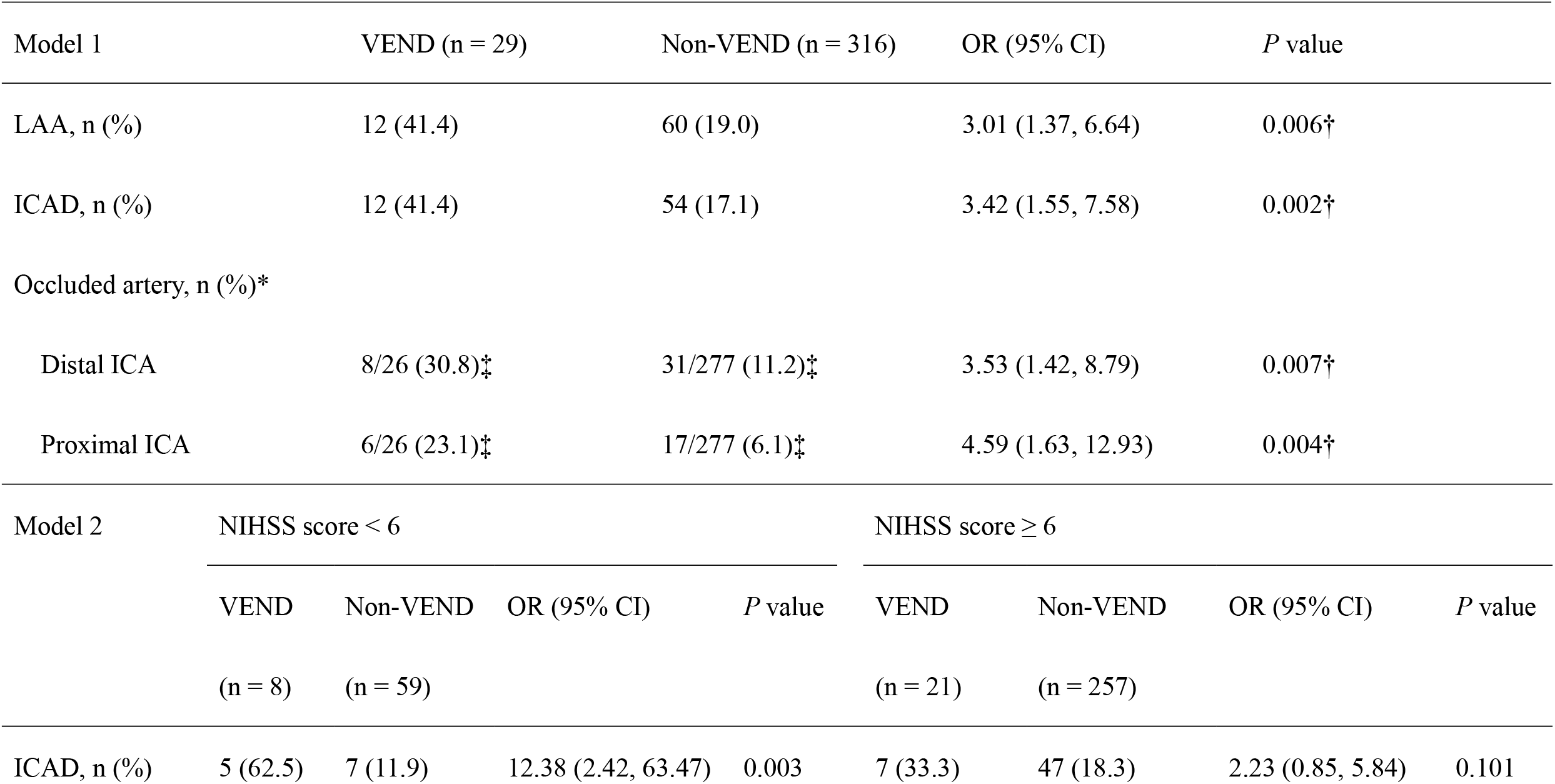
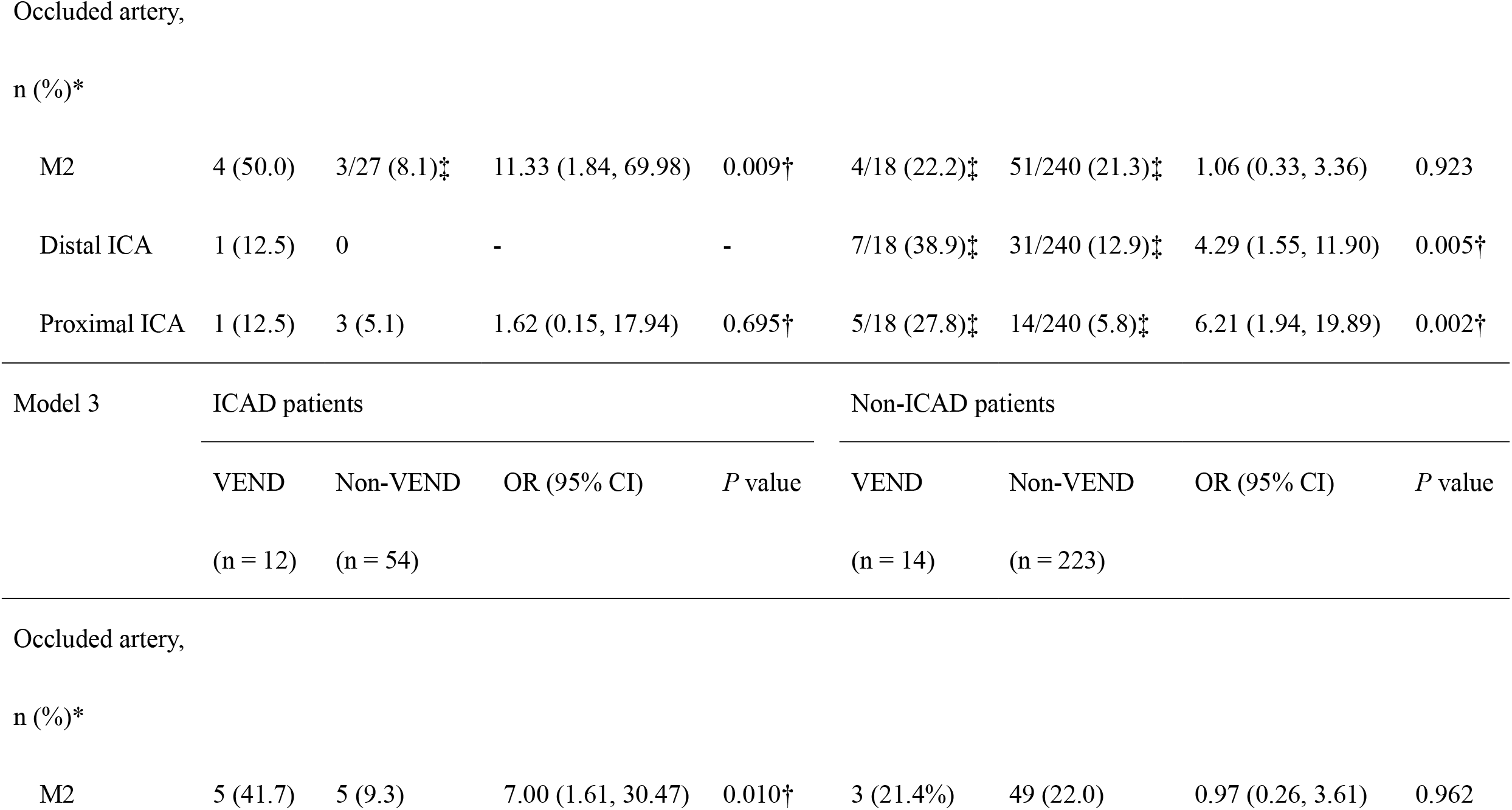
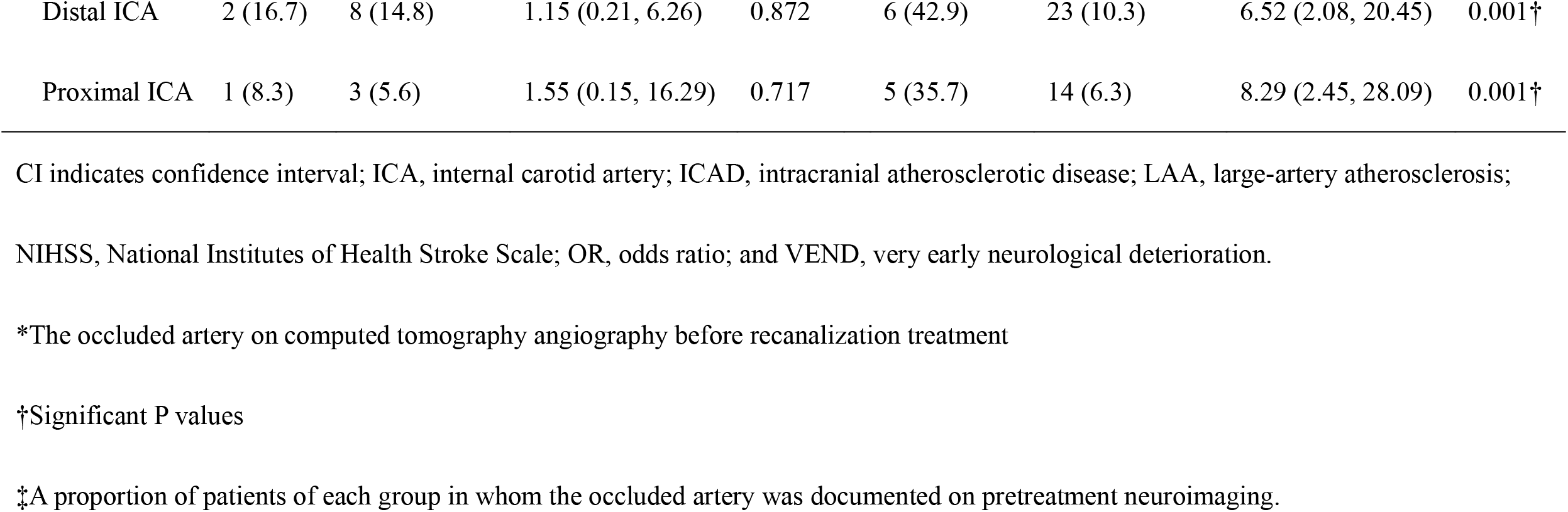
Risk factors significantly associated with VEND in patients receiving thrombolysis

| Model 1 |  | VEND (n = 29) |  | Non-VEND (n = 316) |  | OR (95% CI) |  | P value |  |
| --- | --- | --- | --- | --- | --- | --- | --- | --- | --- |
| LAA, n (%) |  | 12 (41.4) |  | 60 (19.0) |  | 3.01 (1.37, 6.64) |  | 0.006† |  |
| ICAD, n (%) |  | 12 (41.4) |  | 54 (17.1) |  | 3.42 (1.55, 7.58) |  | 0.002† |  |
| Occluded artery, n (%)* |  |  |  |  |  |  |  |  |  |
| Distal ICA |  | 8/26 (30.8)‡ |  | 31/277 (11.2)‡ |  | 3.53 (1.42, 8.79) |  | 0.007† |  |
| Proximal ICA |  | 6/26 (23.1)‡ |  | 17/277 (6.1)‡ |  | 4.59 (1.63, 12.93) |  | 0.004† |  |
| Model 2 |  | NIHSS score < 6 |  |  |  | NIHSS score ≥ 6 |  |  |  |
|  |  | VEND | Non-VEND | OR (95% CI) | P value | VEND | Non-VEND | OR (95% CI) | P value |
|  |  | (n = 8) | (n = 59) |  |  | (n = 21) | (n = 257) |  |  |
| ICAD, n (%) |  | 5 (62.5) | 7 (11.9) | 12.38 (2.42, 63.47) | 0.003 | 7 (33.3) | 47 (18.3) | 2.23 (0.85, 5.84) | 0.101 |

|  |  |  |  |  |  |  |  |  |
| --- | --- | --- | --- | --- | --- | --- | --- | --- |
| M2 | 4 (50.0) | 3/27 (8.1)‡ | 11.33 (1.84, 69.98) | 0.009† | 4/18 (22.2)‡ | 51/240 (21.3)‡ | 1.06 (0.33, 3.36) | 0.923 |
| Distal ICA | 1 (12.5) | 0 | - | - | 7/18 (38.9)‡ | 31/240 (12.9)‡ | 4.29 (1.55, 11.90) | 0.005† |
| Proximal ICA | 1 (12.5) | 3 (5.1) | 1.62 (0.15, 17.94) | 0.695† | 5/18 (27.8)‡ | 14/240 (5.8)‡ | 6.21 (1.94, 19.89) | 0.002† |

| Model 3 | ICAD patients |  |  |  | Non-ICAD patients |  |  |  |
| --- | --- | --- | --- | --- | --- | --- | --- | --- |
|  | VEND | Non-VEND | OR (95% CI) | *P* value | VEND | Non-VEND | OR (95% CI) | *P* value |
|  | (n = 12) | (n = 54) |  |  | (n = 14) | (n = 223) |  |  |

|  |  |  |  |  |  |  |  |  |
| --- | --- | --- | --- | --- | --- | --- | --- | --- |
| M2 | 5 (41.7) | 5 (9.3) | 7.00 (1.61, 30.47) | 0.010† | 3 (21.4%) | 49 (22.0) | 0.97 (0.26, 3.61) | 0.962 |
| Distal ICA | 2 (16.7) | 8 (14.8) | 1.15 (0.21, 6.26) | 0.872 | 6 (42.9) | 23 (10.3) | 6.52 (2.08, 20.45) | 0.001† |
| Proximal ICA | 1 (8.3) | 3 (5.6) | 1.55 (0.15, 16.29) | 0.717 | 5 (35.7) | 14 (6.3) | 8.29 (2.45, 28.09) | 0.001† |

CI indicates confidence interval; ICA, internal carotid artery; ICAD, intracranial atherosclerotic disease; LAA, large-artery atherosclerosis;
NIHSS, National Institutes of Health Stroke Scale; OR, odds ratio; and VEND, very early neurological deterioration.
\*The occluded artery on computed tomography angiography before recanalization treatment
†Significant P values
‡A proportion of patients of each group in whom the occluded artery was documented on pretreatment neuroimaging.

In patients with minor ischemic stroke (NIHSS score < 6) who were not eligible for EVT before deterioration, the occurrence of VEND after thrombolysis was also significantly associated with symptomatic ICAD (OR 12.38, 95% CI 2.42–63.47, *P* = 0.003) (Table 3). Additionally, in the minor stroke group, patients with VEND were more likely to receive rescue EVT (OR 11.20, 95% CI 1.77-70.76, P = 0.019). Among the 304 patients (88.1%) having CTA image before recanalization therapy, pretreatment occlusion in the middle cerebral artery (MCA) M2 segment (OR 11.33, 95% CI 1.84–69.98, *P* = 0.009) instead of M1 segment was associated with VEND (Table 3). Conversely, in patients with an initial NIHSS score of ≥ 6, VEND was associated with pretreatment occlusions in the distal or proximal ICA. Similar to patients with minor stroke, the occurrence of VEND in those with non-minor stroke was associated with higher probability of receiving EVT (OR 4.86, 95% CI 1.88-12.50, *P* = 0.001).

Among the 66 patients with ICAD-related AIS, 12 (18%) suffered from VEND after receiving thrombolysis. In patients with ICAD, VEND was significantly associated with pretreatment M2 occlusion (OR 7.00, 95% CI 1.61–30.47, *P* = 0.010) (Table 2). In contrast, the occurrence of VEND in patients without ICAD was associated with pretreatment occlusion in the distal or proximal ICA.

### Determinants of poor functional outcome in patients with VEND

In patients with VEND, the determinants for 3-month poor functional outcomes were the NIHSS score at 24 h (OR 1.46, 95% CI 1.04–2.06, *P* = 0.031) and the reduced NIHSS score at 24 h compared to that at 1 h (OR 1.17, 95% CI 1.00–1.35, *P* = 0.043), but not the NIHSS scores before treatment or at 1 h after initiating thrombolysis (Table 4). Among patients receiving thrombolysis and EVT, the proportion of patients with an mRS > 2 at 3 months was similar between those with and without VEND (65% vs. 49%, *P* = 0.289).

**Table 4.** Factors associated with poor functional outcome at 3 months in patients with VEND after thrombolysis

|  | OR | 95% CI | *P* value |
| --- | --- | --- | --- |
| Initial NIHSS score | 1.10 | 0.94, 1.28 | 0.249 |
| NIHSS score at 1 h after initiating tPA | 1.12 | 0.97, 1.29 | 0.110 |
| NIHSS score at 24 h after initiating tPA | 1.46 | 1.04, 2.06 | 0.031† |
| Changes of NIHSS scores in 24 h after initiating tPA* | 1.17 | 1.00, 1.35 | 0.043† |

CI indicates confidence interval; h, hour(s); NIHSS, National Institutes of Health Stroke
Scale; OR: odds ratio; and VEND, very early neurological deterioration.
\*NIHSS score at 24 h subtracted by NIHSS score at 1 h after initiating tPA
†Significant *P* values

### Efficacy of EVT in patients with VEND

Given that patients with VEND had higher 24-h NIHSS scores which were significantly associated with poor functional outcomes, we further analyzed the efficacy of EVT on the NIHSS scores at 24 h in these patients (Figure 2). In patients with VEND, those with successful recanalization (mTICI ≥ 2b) by EVT had lower NIHSS scores at 24 h compared to those without successful recanalization (12 ± 9 vs. 26 ± 7, *P* = 0.047). Furthermore, in patients receiving thrombolysis and EVT, there was no significant difference between VEND and non-VEND group in NIHSS scores at 24 h and changes in these scores at 24 h (Supplemental Figure 2).

**Figure 2.**
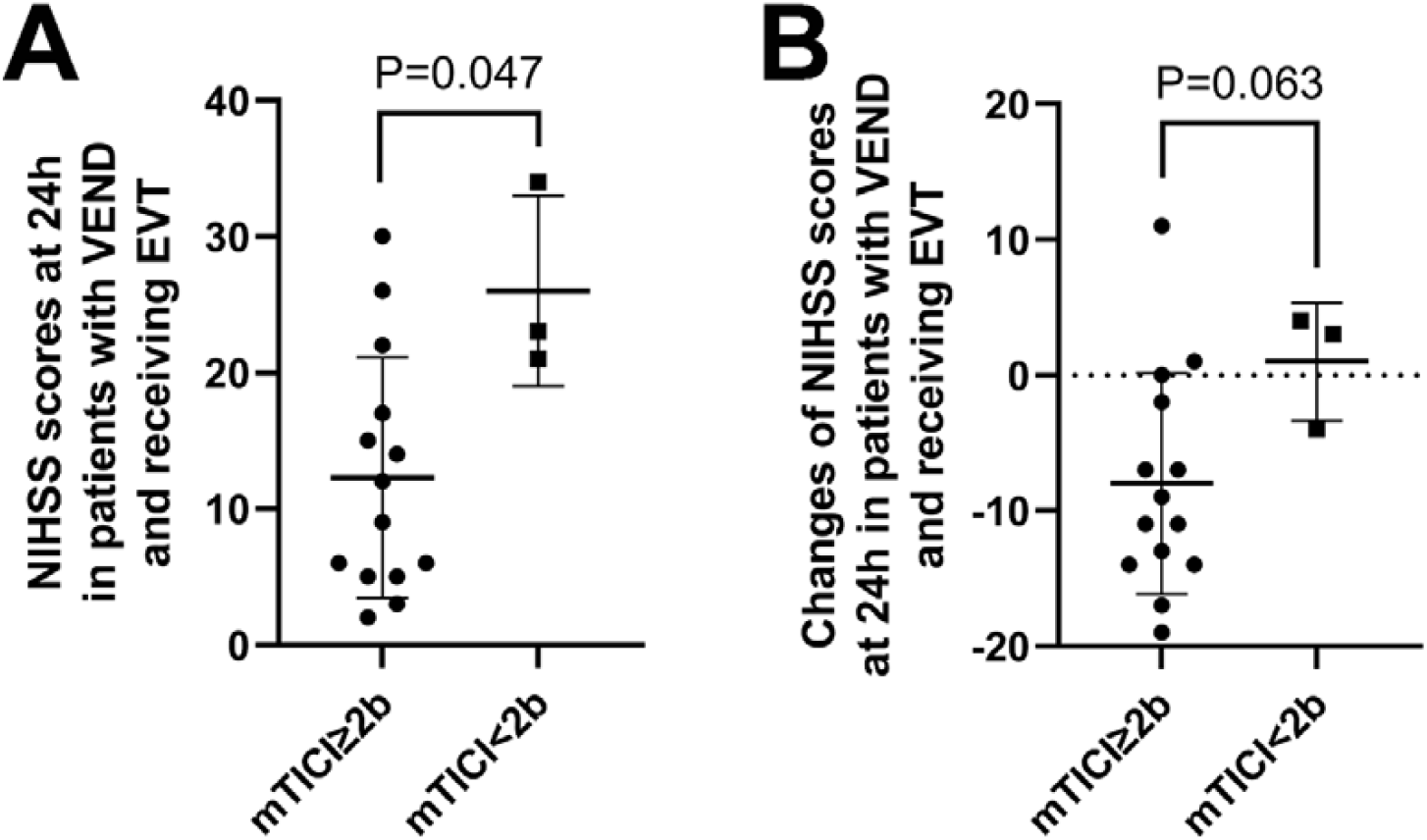
Relationship between successful recanalization and NIHSS score at 24 h in patients receiving EVT after VEND. EVT, endovascular thrombectomy; NIHSS, National Institutes of Health Stroke Scale; VEND, very early neurological deterioration. (A) In patients receiving EVT after VEND, successful recanalization (mTICI ≥ 2b) led to lower NIHSS scores at 24 h compared to those without successful recanalization (12 ± 9 vs. 26 ± 7, *P* = 0.047). (B) There was a trend of greater reduction in NIHSS scores at 24 h (NIHSS scores at 24 h subtracted by those at 1 h) after successful recanalization (−8 ± 8 vs. 1 ± 4, *P* = 0.063). All these comparisons were calculated using Mann-Whitney *U* test. mTICI, modified Thrombolysis in Cerebral Infarction Score.

## Discussion

The present study clarified the prevalence, risk factors, and outcomes of VEND within 1 h of initiating tPA therapy. The rate of VEND in patients receiving thrombolysis was 8.4%. The occurrence of VEND after thrombolysis was significantly associated with symptomatic ICAD. In patients with minor ischemic stroke who were initially not eligible for EVT, exhibiting VEND strongly indicated underlying ICAD, receiving rescue EVT, and it was the only significant factor predicting poor functional outcomes. These findings suggest that ICAD plays a crucial role in the development of VEND after thrombolysis, and intravenous thrombolysis might not be effective in treating patients with ICAD who present with minor stroke. Furthermore, in patients with VEND, successful recanalization by EVT resulted in a lower NIHSS score at 24 h, which significantly predicted the functional outcome at 3 months. Although VEND was an independent predictor of poor functional outcomes at 3 months in patients receiving thrombolysis, the VEND and non-VEND groups did not differ in the 24-h NIHSS scores and 3-month functional outcomes after thrombolysis and EVT, which probably attributed to the efficacy of EVT.

This study showed for the first time that VEND in patients receiving thrombolysis is not rare and that ICAD plays a major role in the pathophysiologic mechanisms of acute neurologic deterioration. Previous reports have shown that patients with LAA have a greater risk of developing END after thrombolysis in AIS.^5,22,23^ Another study found that acute neurological deterioration was observed at a median of 3.6 h from arrival in nearly 20% patients with large-vessel occlusion and mild stroke.^24^ In our study, there was a significant association between the occurrence of VEND and underlying LAA, especially ICAD. We further showed different pretreatment-occluded vessels in the VEND group with minor and non-minor stroke. In patients with minor stroke, pretreatment occlusion at MCA M2 had a higher risk of VEND after thrombolysis. In these patients, partial occlusion before thrombolysis was speculated based on the minor presentation, followed by active thrombus formation induced by exposure of the unstable intima after thrombolysis.^25^ Conversely, occlusion in the ICA with minor stroke did not increase the risk of VEND, probably because tPA could scarcely recanalize the large thrombus occluding the ICA.^26^ Another explanation was that the ICA occlusion was probably chronic and the mechanisms of the minor stroke were acute hemodynamic insufficiency or artery-to-artery embolization, which were less likely to be worsened by thrombolysis.^25,27^ In patients with non-minor stroke, this study showed that pretreatment ICA occlusion was associated with an increased risk of VEND after thrombolysis. In these cases, the ICA occlusion was more likely to be acute occlusion by a large clot, and the high stroke severity indicated inadequate collateral which might be worsened by the reduction of blood pressure before thrombolysis.^27,28^ Therefore, there might be differences in occluded arteries and mechanisms in VEND events of different stroke severities.

This study revealed that VEND had a significant detrimental impact on 3-month functional outcomes, in concordance with previous findings for END.^29,30^ Of note, this study further revealed that VEND significantly predicted poor outcomes in both minor and non-minor stroke patients. A prior study showed that 12% of patients who received thrombolysis therapy for acute minor ischemic stroke due to large vessel occlusion developed ischemic END and 48% of the END occurred within 2 h, which was associated with worse 3-month functional outcomes even after rescue thrombectomy.^30^ However, our study showed that (1) early successful recanalization by EVT in patients with VEND significantly correlated with lower NIHSS scores at 24 h, and (2) there was no difference in the NIHSS scores at 24 h and in functional outcomes 3 months after EVT between the VEND and non-VEND groups. These findings demonstrated the importance of rescue thrombectomy with successful recanalization in patients with VEND after thrombolysis.

This study has several limitations. First, our research was a registry-based retrospective study; thus, the influence of missing data and selection bias should be taken into consideration. Second, blood pressure before thrombolysis was not recorded continuously, so the range of blood pressure reduction before thrombolysis might be underestimated. Third, for patients who were undergoing EVT at 1 h of initiating thrombolysis, underestimation of VEND during EVT procedure could not be excluded especially for those with impaired consciousness or under general anesthesia. However, only 4% of our patients received general anesthesia for EVT procedure, which might minimize the likelihood of underestimating VEND during EVT.

## Conclusion

In patients with AIS receiving intravenous thrombolysis, VEND was an independent predictor of poor functional outcomes at 3 months, which was especially important for those with minor stroke and underlying ICAD. For patients with VEND, successful recanalization by EVT led to a lower stroke severity at 24 h, which associated with long-term functional outcome. These findings emphasized the importance of identifying VEND in patients receiving thrombolysis due to its independent association with poor functional outcomes and benefit of successful recanalization by rescue EVT if large-vessel occlusion is revealed.

## Data Availability

Data available on request from the authors.

## Acknowledgements

Not applicable.

## Supplemental Material

Supplemental Figure 1

Supplemental Figure 2

Supplemental Table 1

## Non-standard Abbreviations and Acronyms

AIS: acute ischemic stroke
AUC: area under the ROC curve
aOR: adjusted odds ratio
CI: confidence interval
CTA: computed tomography angiography
DBP: diastolic blood pressure
ECAD: extracranial atherosclerotic disease
END: early neurological deterioration
EVT: endovascular thrombectomy
HbA1c: hemoglobin A1C
ICA: internal carotid artery
ICAD: intracranial atherosclerotic disease
INR: international normalized ratio
LAA: large artery atherosclerosis
LDL: low-density lipoprotein cholesterol
mRS: modified Rankin Scale
mTICI: modified Thrombolysis in Cerebral Infarction Score
NIHSS: National Institutes of Health Stroke Scale
OR: odd ratio
SBP: systolic blood pressure
sICH: symptomatic intracerebral hemorrhage
ROC: receiver operating characteristic
TG: triglycerides
tPA: tissue plasminogen activator
VEND: very early neurological deterioration

